# A comparison of large language model versus manual chart review for extraction of data elements from the electronic health record

**DOI:** 10.1101/2023.08.31.23294924

**Authors:** Jin Ge, Michael Li, Molly B. Delk, Jennifer C. Lai

**Affiliations:** Division of Gastroenterology and Hepatology, Department of Medicine, University of California – San Francisco, San Francisco, CA; Section of Gastroenterology and Hepatology, Department of Medicine, Tulane University School of Medicine, New Orleans, LA

**Author notes:** **<u>Correspondence:</u>** Jin Ge, MD, MBA, 513 Parnassus Avenue, S-357, San Francisco, CA 94143. **<u>Writing Assistance:</u>** None. **<u>Author Contributions:</u>** Authorship was determined using ICMJE recommendations. *Ge:* Study concept and design; data extraction; analysis and interpretation of data; drafting of manuscript; critical revision of the manuscript for important intellectual content; statistical analysis; study supervision *Li:* Analysis and interpretation of data; data extraction; critical revision of the manuscript for important intellectual content *Delk:* Analysis and interpretation of data; data extraction; critical revision of the manuscript for important intellectual content *Lai:* Study concept and design; analysis and interpretation of data; critical revision of the manuscript for important intellectual content; study supervision.

## Abstract

**Importance:** Large language models (LLMs) have proven useful for extracting data from publicly available sources, but their uses in clinical settings and with clinical data are unknown.

**Objective:** To determine the accuracy of data extraction using “Versa Chat,” a chat implementation of the general-purpose OpenAI gpt-35-turbo LLM model, versus manual chart review for hepatocellular carcinoma (HCC) imaging reports.

**Design:** We engineered a prompt for the data extraction task of six distinct data elements and input 182 abdominal imaging reports that were also manually tagged. We evaluated performance by calculating accuracy, precision, recall, and F1 scores.

**Setting/Participants:** Cross-sectional abdominal imaging reports of patients diagnosed with hepatocellular carcinoma enrolled in the Functional Assessment in Liver Transplantation (FrAILT) study.

## Background

Large language models (LLMs) hold tremendous potential for accelerating clinical research and augmenting clinical care.^1^ One of the most promising LLM use cases is natural language processing (NLP) and extraction of structured elements from unstructured clinical text, such as imaging reports.^2^ LI-RADS (Liver Imaging Reporting and Data System) was created by the American College of Radiology and provides standardized and reproducible reporting of hepatocellular carcinoma (HCC) imaging for clinical care and research.^3^ Due to the LI-RADS reporting system, HCC imaging provides an ideal test case for LLM-enabled NLP extraction of structured data from unstructured clinical text. We sought to assess the performance of a commercially available general-purpose LLM, deployed in an isolated protected environment and permitted to be used with protected health information (PHI), versus human manual chart review in extracting six distinct data elements from abdominal imaging reports.

## Methods

We used”Versa Chat,” the chat user interface of the general purpose Microsoft Azure OpenAI gpt-35-turbo LLM model (“Versa”) that is implemented in a protected environment at the University of California, San Francisco (UCSF) to accommodate the use of PHI and intellectual property, for this study.^4^”Versa,” like other gpt-35-turbo implementations, has a token limit of 4,096 tokens, defined as the unit that OpenAI generative artificial intelligence (GAI) models use to compute text length. One token approximates to about four characters or one word. This 4,096 token limit includes the count from both the user prompt and completion of the task for each session.^5^ We manually reviewed 182 CT or MRI abdomen imaging reports without evidence of locoregional treatments from 169 patients diagnosed with HCC enrolled in the Functional Assessment in Liver Transplantation (FrAILT) study at UCSF.^6^ The imaging reports, therefore, may or may not contain evidence of HCC as a diagnosis could have occurred subsequent to the date of imaging. We manually tagged the imaging reports for six distinct data elements: 1. Maximum LI-RADS score for any HCC lesions (defined as 4 or 5), 2. Number of HCC lesions, 3. Diameter (cm) of the largest lesion, 4. Sum of diameters (cm) of all HCC lesions, 5. Presence or absence of macrovascular invasion, or 6. Presence or absence of extrahepatic metastases.

All 182 imaging reports were trimmed to only include the findings and impressions sections. Due to the limitation of 4,096 tokens per session in”Versa Chat,” we iteratively developed a”zero-shot” prompt (defined as a prompt that does not contain training data) with testing on the first five records (Figure 1).^7^ As snowballing of data passed per prompt often led to execution failure from exceeding the token limit, we ran 26 sessions of the final”zero-shot” extraction prompt in”Versa Chat” with approximately seven records per session for data extraction (see Figure 2 for an example exchange using mock data with”Versa Chat”). If”Versa Chat” produced an output that required minor additional formatting, we made those changes within the chat interface prior to collecting and aggregating the data. The total amount of time required to process all 182 records was 45 minutes.

**Figure 1.**
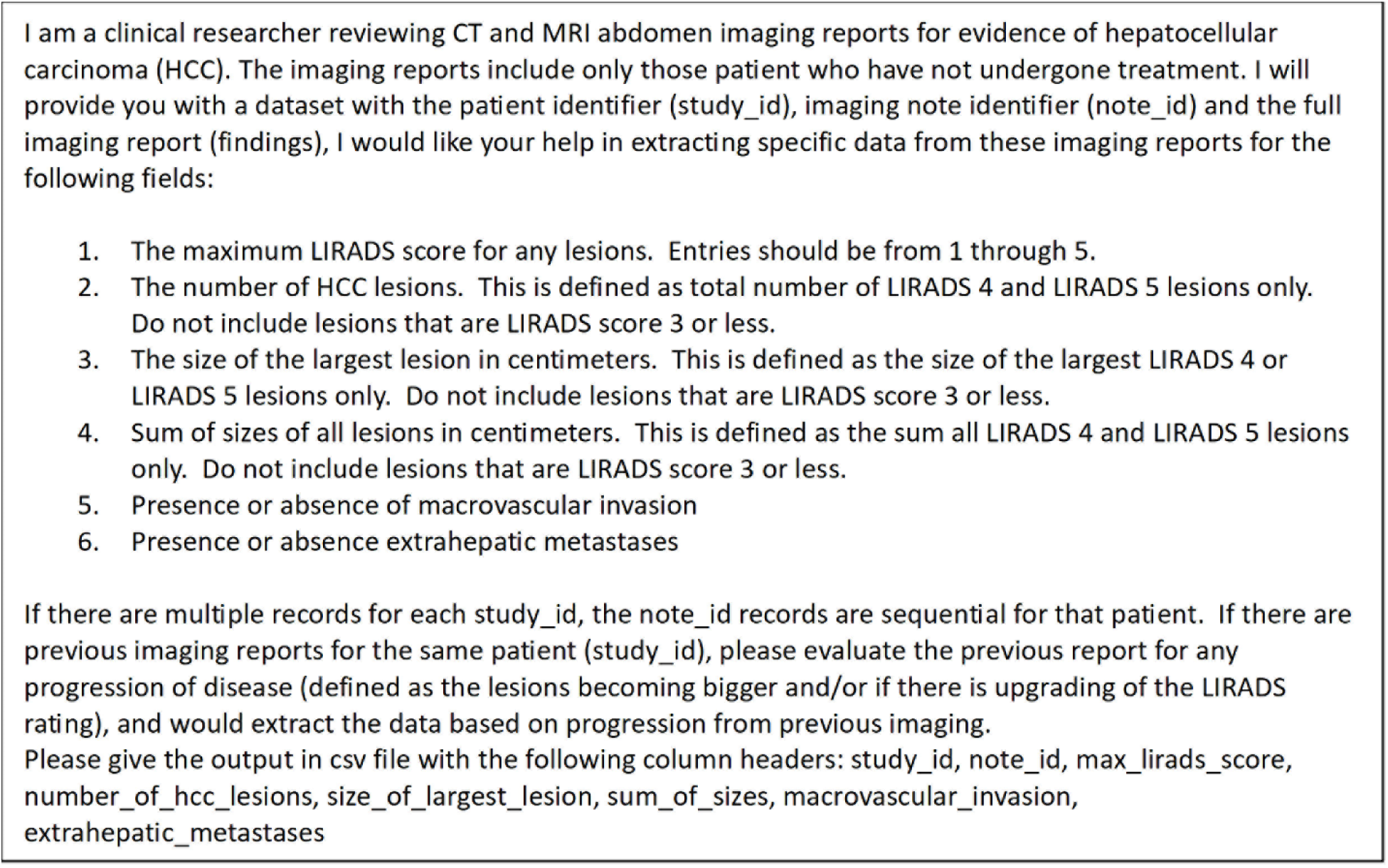
Final prompt used for data extraction from”Versa Chat” (gpt-35-turbo)

**Figure 2.**
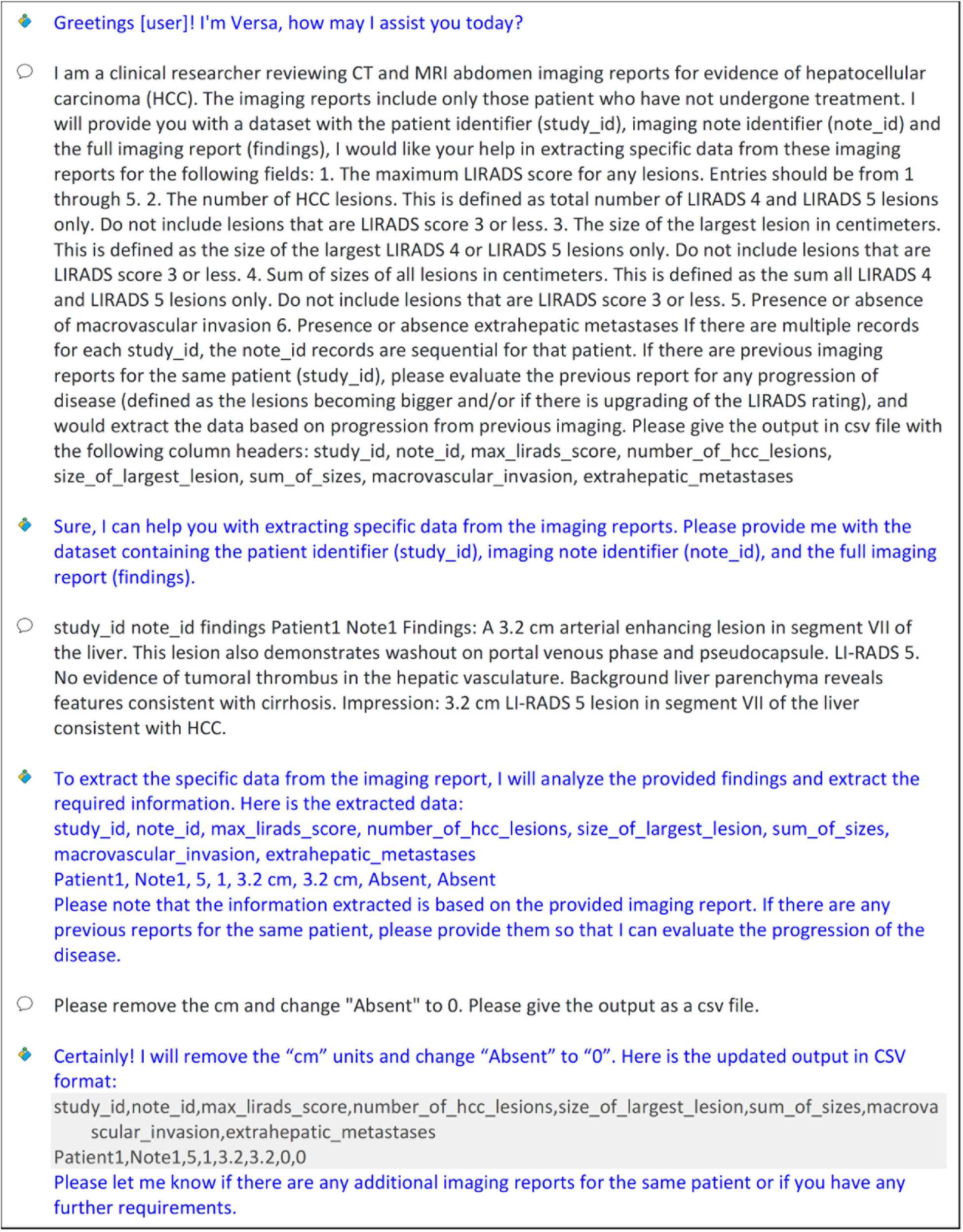
Example of an exchange with”Versa Chat” (gpt-35-turbo) using mock data.

We evaluated the accuracy of”Versa Chat” data extractions versus manual chart review with each imaging report as a separate record. We calculated performance metrics, notably accuracy, precision, recall, and F1 score (harmonic mean of precision and recall commonly used to evaluate classification in machine learning) for each of the six data elements whenever possible. For multilevel classifications (maximum LI-RADS score, number of HCC lesions, diameter of the largest lesion, and sum of tumor diameters), we calculated weighted-average precision, recall, and F1 score. For binary classifications (macrovascular invasion and extrahepatic metastases), we defined the presence of these features as a positive case for precision, recall, and F1 score.^8^ We estimated 95% confidence intervals (CI) for performance metrics whenever possible through bootstrapping with 2,000 iterations. All statistical analyses were conducted in R, version 4.3.1”Beagle Scouts” (R Core Team, Vienna, Austria),^9^ and R packages *boot*, version 1.3-28.1,^10^ and *caret*, version 6.0-94.^11^ This study was approved by the UCSF Institutional Review Board in Study #11-07513.

## Results

The performance metrics for the six data elements extracted by the gpt-35-turbo”Versa Chat” model versus manual chart review are featured in Table 1. The overall accuracy of”Versa Chat” was 0.889 (95% CI 0.869-0.907) versus manual review. The accuracy rate varied between 0.725 (95% CI 0.643-0.780) for sum of tumor diameters to 0.989 (95% CI 0.956-0.995) for macrovascular invasion. In general, accuracy was higher for simple classification tasks (maximum LI-RADS score, macrovascular invasion, and extrahepatic metastases) compared to those that required comparison (maximum tumor diameter) or summation (number of tumors and sum of tumor diameters). As macrovascular invasion and extrahepatic metastases did not have any true positive cases, the precision for these two data elements were both zero. Similarly, as there were no false negative cases, the recall and F1 score for macrovascular invasion could not be calculated. As the precision, recall, and F1 score statistics for maximum LI-RADS score, number of tumors, maximum tumor diameter, and sum of tumor diameters were calculated as weighted-average values due to multilevel classifications, these values may be biased as accurate predictions of absence of an imaging feature (e.g.”Versa Chat” noted zero tumors when there were no tumors by manual chart review) were included in the statistics.

**Table 1.**
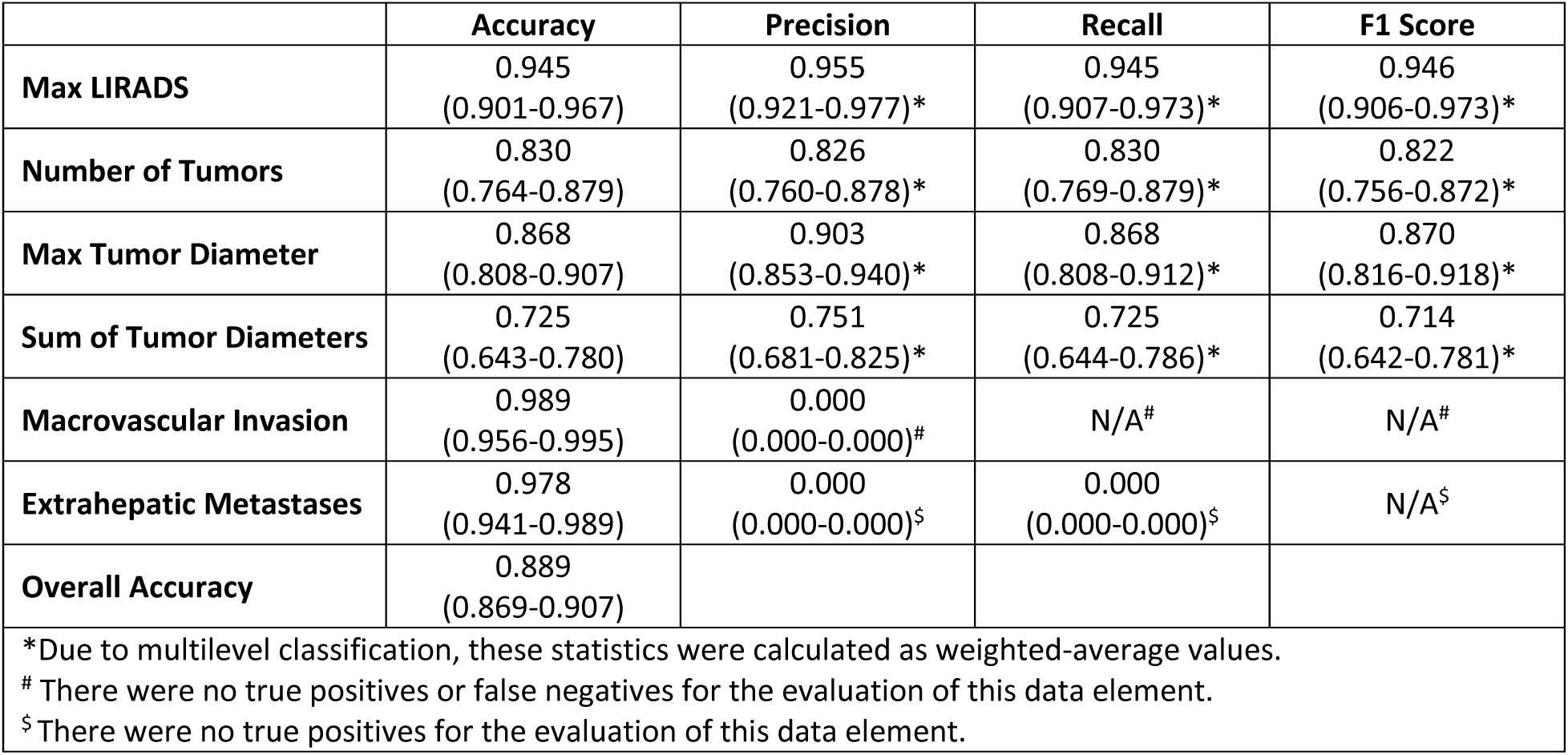
Performance evaluation statistics of”Versa Chat” versus manual chart review.

|  | **Accuracy** | **Precision** | **Recall** | **F1 Score** |
| --- | --- | --- | --- | --- |
| **Max LIRADS** | 0.945<br>(0.901-0.967) | 0.955<br>(0.921-0.977)* | 0.945<br>(0.907-0.973)* | 0.946<br>(0.906-0.973)* |
| **Number of Tumors** | 0.830<br>(0.764-0.879) | 0.826<br>(0.760-0.878)* | 0.830<br>(0.769-0.879)* | 0.822<br>(0.756-0.872)* |
| **Max Tumor Diameter** | 0.868<br>(0.808-0.907) | 0.903<br>(0.853-0.940)* | 0.868<br>(0.808-0.912)* | 0.870<br>(0.816-0.918)* |
| **Sum of Tumor Diameters** | 0.725<br>(0.643-0.780) | 0.751<br>(0.681-0.825)* | 0.725<br>(0.644-0.786)* | 0.714<br>(0.642-0.781)* |
| **Macrovascular Invasion** | 0.989<br>(0.956-0.995) | 0.000<br>(0.000-0.000)# | N/A# | N/A# |
| **Extrahepatic Metastases** | 0.978<br>(0.941-0.989) | 0.000<br>(0.000-0.000)\$ | 0.000<br>(0.000-0.000)\$ | N/A\$ |
| **Overall Accuracy** | 0.889<br>(0.869-0.907) |  |  |  |

\*Due to multilevel classification, these statistics were calculated as weighted-average values.

## Discussion

This is one of the first studies that has demonstrated and compared the performance of the chat interface of a general-purpose LLM versus manual chart review for extraction of clinical data. We demonstrated high accuracy for simple extraction tasks, which degraded with more complex use cases. Of note, iterative development (“prompt engineering”) of a”zero-shot” prompt to specify the operations to be executed by the LLM was necessary to achieve this level of accuracy.^7^ Our use of a”zero-shot” prompt and limiting the amount of data processed per session, however, prevented the gpt-35-turbo model from maintaining a persistent memory to allow in-context”learning” based on previous data.^12^ These are known limitations of the gpt-35-turbo model, which have been improved upon in gpt-35-turbo-16k (which supports up to 16,384 tokens), gpt-4 (up to 8,192 tokens), and gpt-4-32k (up to 32,768 tokens).^5^ Despite these limitations, our study demonstrated two important concepts: 1. Feasibility of using general purpose LLMs to extract structured information from clinical data with *minimal* technical expertise, and 2. Use of a LLM deployed in isolated protected environment that accommodates PHI (as opposed to ChatGPT, which is often not permitted for use with PHI) for clinical use cases.

## Data Availability

Aggregate data produced in the present study may be available upon reasonable request to and with approval by the authors.

## Abbreviations

CI: confidence interval
FrAILT: Functional Assessment in Liver Transplantation
GAI: generative artificial intelligence
HCC: hepatocellular carcinoma
LI-RADS: Liver Imaging Reporting and Data System
LLM: large language model
NLP: natural language processing
PHI: protected health information
UCSF: University of California, San Francisco

## Data Acknowledgement

- The authors thank the UCSF AI Tiger Team, Academic Research Services, Research Information Technology, and the Chancellor’s Task Force for Generative AI for their software development, analytical, and technical support related to the use of Versa API gateway (the UCSF secure implementation of large language models and generative AI via API gateway), Versa chat (the chat user interface), and related data assets.

